# Mental health self-care during the COVID-19 pandemic: A prospective cohort study in Australia

**DOI:** 10.1101/2022.12.08.22283265

**Authors:** Daniel Griffiths, Vinsensia Maharani Kanya Dhira Pradipta, Alex Collie

## Abstract

Pandemic public health measures have affected mental health for many people, resulting in varied approaches to mental health self-care. During 27 April – 26 July 2020, we surveyed a cohort of 1646 Australians, who were in paid employment prior to the pandemic, on changes in work, health, and managing their mental health concerns. Lifestyle changes were most the most frequently reported action to manage mental health concerns (78%), and were more common for women (OR=2.33, 95%CI=[1.82, 3.03]), and people experiencing recent work loss (OR=1.54, 95%CI=[1.04, 2.28]). Mental health self-care was more common for people experiencing psychological distress, or with pre-exisiting mental health conditions. Talking to friends about mental health, exercise and dietary changes, were more common for women and younger adults. Findings highlight potential benefits of reducing barriers to formal mental health services and supports during crises, particularly for people who less commonly seek help, and those experiencing psychological distress.

## Introduction

Large increases in mental health problems including depression and anxiety symptoms, and psychological stress, have been highlighted from the early stages of the COVID-19 pandemic and during periods of restrictions. In response, many developed countries established dedicated telephone support lines or online mental health advice information for people in need of support, and some governments increased mental health funding and access to mental health services (OECD 2021). Mental health support organisations have also promoted actions that individuals could take to support their mental health such as staying active and socially connected, and discouraged potentially harmful actions such as consuming alcohol or illicit drugs. Whilst information on formal mental health service use is routinely captured by healthcare administrative systems (e.g. for billing purposes) and reported by Government health departments, other behavioural approaches to promote resilience and wellbeing, or informal mental health supports like talking to friends or family, are typically unreported.

Self-care is the ability of people promote and maintain health, and cope with illness, either with or without the support of a health care provider (WHO 2014). Several approaches to mental health self-care pivoted during the COVID-19 pandemic to align with periods of public health restrictions. For example, telehealth services acted as a substitute for in-person health consultations (Fisk, Livingstone, and Pit 2020), and in-person group social activities pivoted to virtual forms in some circumstances. During periods of restrictions and lockdowns there was increased focus on forms of self-care such as practicing meditation and mindfulness at home (Deguma et al. 2021), limiting news and social media exposure (Gao et al. 2020; Stainback, Hearne and Trieu 2020), involvement in distraction activities (such as exercise, music or online yoga) and communicating with family, friends and colleagues about personal mental health concerns (Labrague 2021). Whilst the pandemic has led to new mental health promotion information recommending actions to manage mental health concerns, it is less clear to what extent particular forms of mental health self-care were adopted.

The mental health impacts of the COVID-19 pandemic are not shared equally. Some people are more at-risk of ill mental health than others, in part, due to the disruption of several social determinants of health such as pandemic-related loss of work or income (Griffiths et al. 2022b). Those with pre-existing mental health conditions (Griffiths et al. 2021) may also be at increased risk, and risk of mental health problems changes during lockdowns (Griffiths et al. 2022a). As a result, engagement with mental health services, or other forms of self-care may be more relevant, and common, for people experiencing psychological distress, particularly for people with ongoing counselling or prescribed with medications for pre-existing mental health conditions. Barriers to mental health help-seeking behaviour are known to differ by gender, with men less commonly utilising health services, reportedly due to personal stigma and increased stoicism (Judd, Komiti and Jackson 2008), and older adults are known to be especially unlikely to seek mental health services despite a positive attitude to such services (Mackenzie et al. 2008). By understanding which forms of mental health self-care were less commonly undertaken during the COVID-19 pandemic by specific subgroups, it is possible to identify specific populations who may benefit from helpful forms of mental health self-care, whilst discouraging potentially harmful actions.

Using a prospective cohort of working-age Australians enrolled in a study of changes in health and work during the first year of the COVID-19 pandemic, we sought to answer two research questions on actions taken to manage mental health concerns during the pandemic.

1. How prevalent were actions to manage mental health concerns?
2. What demographic, health and employment characteristics were associated with actions taken to manage mental health concerns?

## Methods

### Study design and setting

The COVID-19 Work and Health Study is a national prospective longitudinal cohort study of working-age Australians during 2020 and 2021 (Griffiths et al. 2021). Respondents were all engaged in paid work prior to the COVID-19 pandemic, aged 18 years or older and resided in Australia. Upon study enrolment, respondents completed a 20-minute baseline survey including changes in work, health, pre-existing health conditions and demographics. Respondents were then asked to provide consent to be contacted for follow-up surveys, the first being one month after the baseline survey. Approval to conduct the study was provided by Monash University Human Research Ethics Committee (#24003). Informed consent was obtained from all patients for being included in the study. Participation was voluntary.

### Outcomes

#### Actions to manage mental health concerns

Outcomes were derived from 13 response items to the following question:

*“Thinking about the past month, which of the following actions have you taken to help manage stress, anxiety or other mental health problems?”*. Response items were developed by the study investigator team to reflect both actions promoted by mental health and wellbeing support organisations during the pandemic such as Beyond Blue (Beyond Blue 2022) as a primary reference, as well as the use of dedicated COVID-19 telephone support lines (AusGov DoH 2022), and response items also reflected issues being raised in public discussions of mental health including potentially harmful actions such as reported increases in alcohol consumption (Ramalho 2020).

Responses were collected as binary yes/no variables from 13 response items. These individual response items were allocated to one of five categories: (1) Lifestyle changes, (2) Diet and exercise, (3) Alcohol or drug use, (4) Medications, (5) Helplines and online resources. Outcomes describe whether individuals reported at least one action within any given category, or not.

#### Talking to others about mental health concerns

Respondents were asked the following question: “*In the past month, have you spoken with any of the following people specifically about your mental health?”*. Response options included talking to a friend, family member, general practitioner (GP), psychologist, psychiatrist, another health professional, or someone else (specified). For each respondent, three binary variables were defined as having spoken to at least one type of (1) health professional, or (2) non-health professional about mental health, or (3) having spoken to no one.

#### Predictors

Factors known to be associated with mental health were included as predictors for analyses including gender, age group, employment, recent engagement in work, pre-existing health conditions (anxiety and/or depression), current level of psychological distress, and survey mode (online or telephone) (Griffiths 2021). Covariate reference groups were selected to align with groups of lower psychological distress levels.

Data on pre-existing mental health conditions such as anxiety and depression were collected at baseline. During a 1-month follow-up survey, the 6-item Kessler Psychological Distress (K-6) scale (Kessler et al. 2002) evaluated the current level of distress. Participants were categorised into two groups (1) moderate to high distress: scores in the interval 11 to 30, and (2) low to no distress: scores in the interval 6 to 10 (Kessler et al 2003).

Current details of work described employment status (either currently employed or unemployed), and engagement in work defined as whether people had worked any hours during the prior week or not.

#### Data analysis

Cohort counts and percentages describe the prevalence of mental health actions across each of five categories, in addition to whether people had either spoken about their mental health to a non-health professional, a health professional, or no one during the prior month.

Eight binary logistic regression models were evaluated across: (1) each of the five outcomes describing categories of actions to manage mental health concerns, and (2) three outcomes on talking about personal mental health concerns with others. Model estimates are described as adjusted odds ratios (AOR) with 95% confidence intervals (CI) and a significance level of 0.05.

## Results

Amongst 1646 respondents, 1445 (88%) had taken at least one action to manage their mental health concerns during the prior month (Table 1). A total of 914 (56%) had spoken to someone specifically about their mental health during the prior month such as a health professional (19%), or a family member or friend (52%). Lifestyle changes were the most common way that people were managing their mental health and was reported by 1276 people (78%). Implementing changes to diet and exercise to manage mental health was the second most common category to mental health concerns (47%), followed by increased alcohol or drug use (27%), medication (22%), and use of helplines or other online resources (20%).

**Table 1.** Prevalence of actions taken to manage personal mental health concerns during the COVID-19 pandemic.

|  | Action to manage mental health by workers N (%) |
| --- | --- |
| **Actions to manage personal mental health concerns** |  |
| *Lifestyle changes* | **1276 (77.5)** |
| 1. Watched or read something uplifting | 849 (51.6) |
| 2. Distracted yourself by keeping active or learning a new skill | 826 (50.2) |
| 3. Reduced your exposure to news and social media | 572 (34.8) |
| 4. Practiced meditation or mindfulness | 466 (28.3) |
| *Diet and exercise* | **772 (46.9)** |
| 5. Made changes to your exercise | 667 (40.5) |
| 6. Made changes to your diet | 435 (26.4) |
| *Alcohol of drug use* | **445 (27.0)** |
| 7. Drunk more alcohol than normal | 355 (21.6) |
| 8. Taken non-prescribed drugs | 134 (8.1) |
| *Medications* | **358 (21.7)** |
| 9. Taken your prescribed medication | 302 (18.3) |
| 10. Made changes to your medications | 132 (8.0) |
| *Helplines and online resources* | **331 (20.1)** |
| 11. Searched for information on managing mental health problems (e.g. on the internet) | 225 (13.7) |
| 12. Participated in an online forum / chat group | 141 (8.6) |
| 13. Called a telephone support line | 42 (2.6) |
| **Speaking to others specifically about your mental health** |  |
| 1. Health professional (e.g. general practitioner, psychologist, psychiatrist, ...) | 306 (18.6) |
| 2. Non-health professional (e.g. friend, family member, ...) | 853 (51.8) |
| 3. Spoken to no one | 732 (44.5) |
| **Total sample of working-age Australians** | **1646 (100.0)** |

Overall, women and younger people more commonly reported helpful actions to manage mental health concerns compared to reference groups, as well as those experiencing loss of work, psychological distress and those with pre-existing anxiety or depression (Figure 1, Figure 2, Appendix A-B).

**Figure 1.**
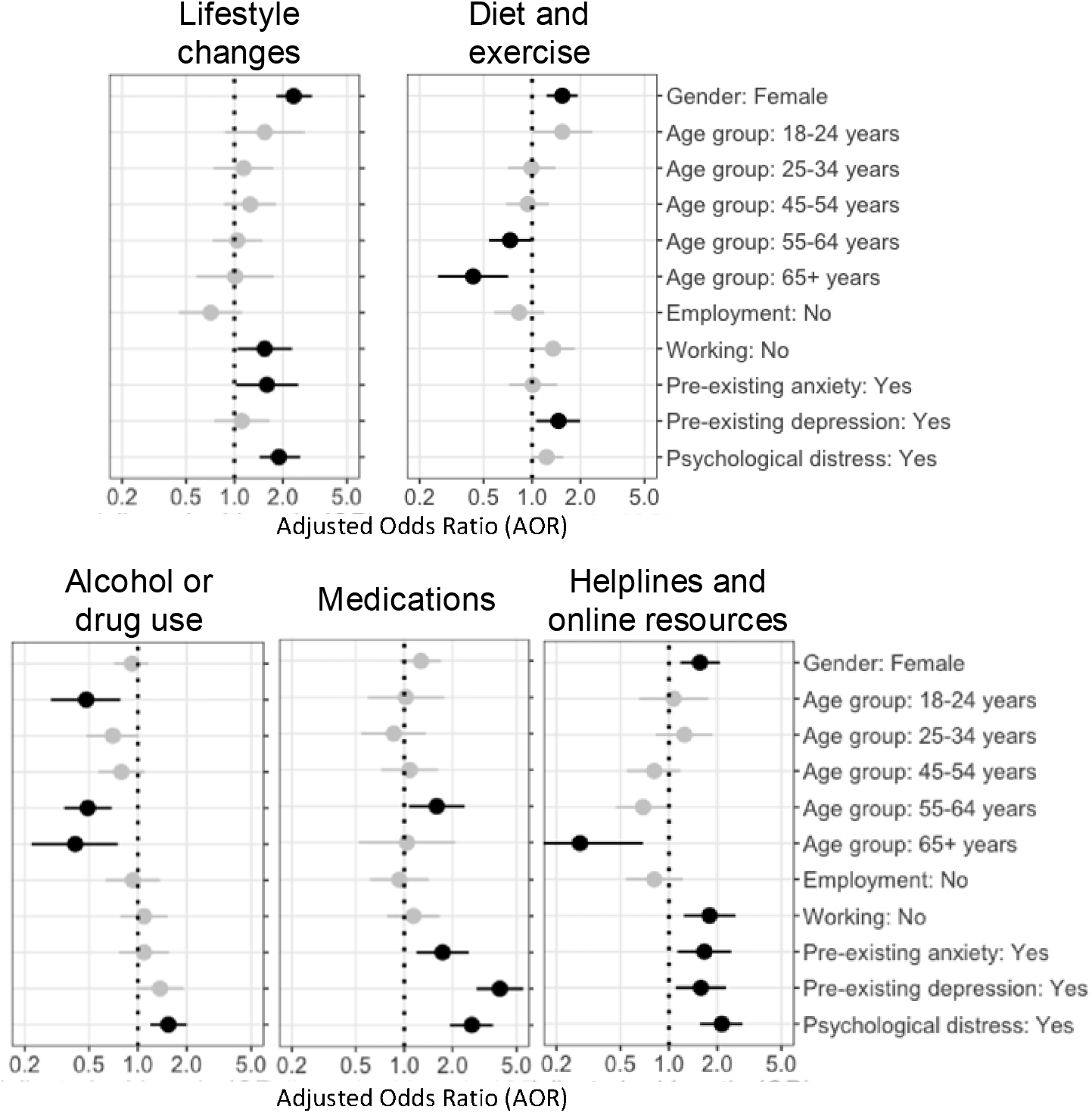
Indicators for taking actions to manage mental health concerns across each of five thematic groups.

**Figure 2.**
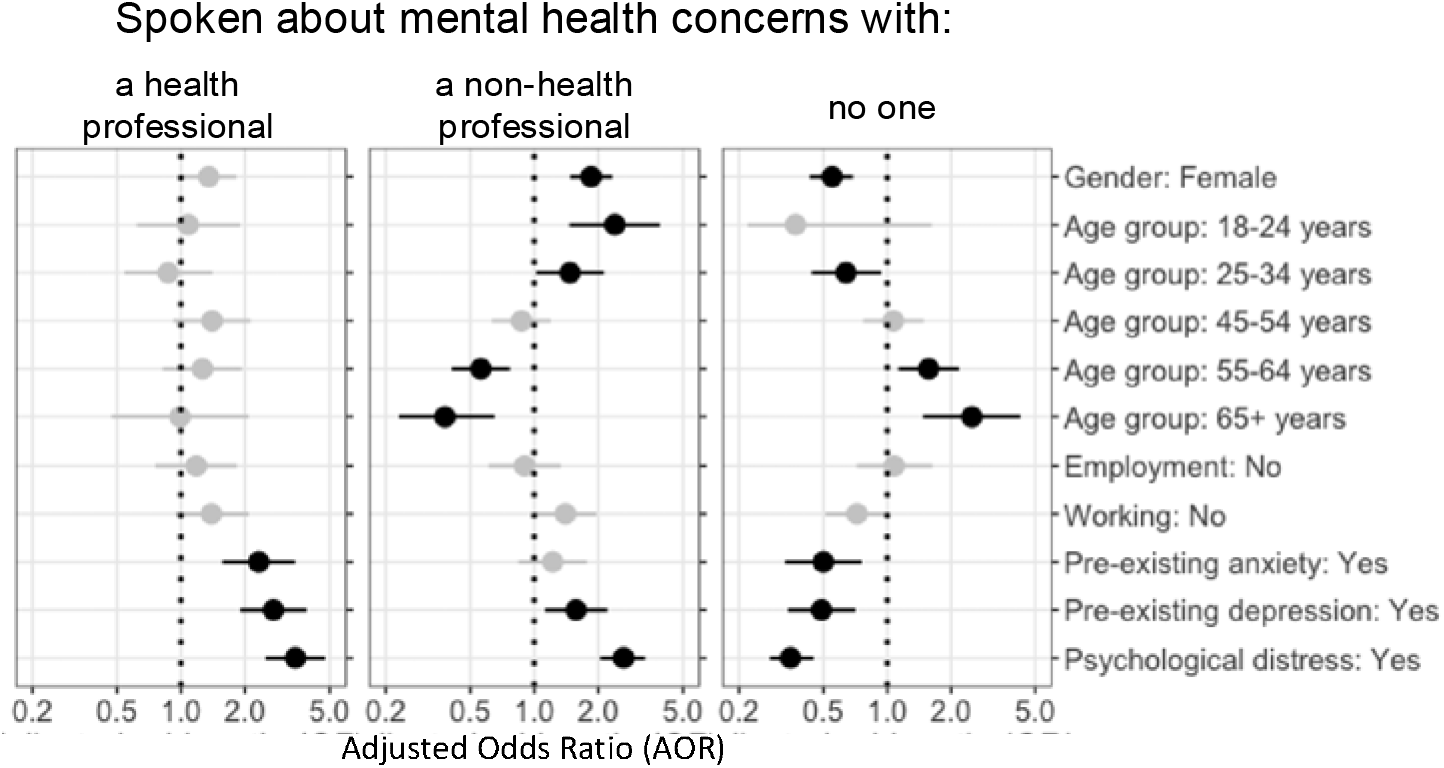
Indicators for talking to others about personal mental health concerns.

Women were more likely to report making changes to their lifestyle, exercise or diet, as well as using online resources or helplines, or talking about their mental health to others such as a family member or friend. Whilst recognising that people over 65 years were relatively uncommon in our cohort (Appendix C), people aged over 55 years were less likely to report making changes to manage their mental health to their exercise, diet, alcohol or drug use, or talking to a friend or family member about their mental health. Increased alcohol consumption or drug use was less prevalent for people aged 18 to 24 years compared to people aged 34 to 44 years. Those aged 55 to 64 years more commonly reported making changes to, or taking, prescribed medications compared to the reference group, and people aged 65 or older less commonly reported using online mental health resources or telephone helplines. People who had not worked during the prior week made changes to their lifestyle and used online resources more often than people who had worked during the prior week.

People experiencing psychological distress, or with pre-existing anxiety or depression, more commonly reported talking to a health professional, using medication to manage their mental health, or searching for help online.

## Discussion

Our findings demonstrate that informal forms of mental health self-care during the COVID-19 pandemic were much more common than the use of formal mental health services such as psychologist or psychiatrist consultations and calls to COVID-19 telephone support lines. Most people reported at least one form of mental health self-care such as meditation or mindfulness, activities to distract themselves, or speaking to a family member or friend about their mental health concerns during the prior month. Despite the heightened use of telephone helplines during the early stages of the pandemic (LifeLine 2020), this was the least common action reported to manage mental health concerns within our sample of working-age Australians. Our findings broadly align with the WHO framework for organisation of services for mental health, describing a higher prevalence of informal service use compared to formal service use (WHO 2003), and our analyses add further value by quantifying more specific informal actions taken to manage mental health. Furthermore, our findings clearly demonstrate the extent to which strategies to manage mental health concerns differ by individuals’ pre-existing and current mental health status, employment, age and gender. The prevalence of actions taken by people in distress suggests that those in most need of help made changes to their lives to support their mental health, and were more likely to seek help. However, people experiencing distress also more commonly increased their consumption of alcohol or drug use, which can lead to poorer health or may contribute towards harmful forms of sustance abuse in some circumstances (Callinan et al. 2021).

Physical health and mental health are closely interrelated, and exercise was commonly reported as an approach to manage mental health concerns during the pandemic. For some people physical activity during the pandemic acted as a motivator for anxiety relief, whereas those with mental health deteriorations have reported increased sedentary behaviour overall (WHO 2003). Physical health interventions, that result in long duration physical activity, may offer the most mental health benefits to men (Bhui and Fletcher 2000), who were identified as having potential barriers to mental health self-care in terms of making changes to their exercise or diet. Furthermore, men may also benefit from employer-based education on mental health supports to reduce negative attitudes of psychological openness to tackle the under-utilisation of mental health services (Mackenzie, Gekoski and Knox 2006), and to benefit from the mental health support of family and friends (Li and Xu 2020).

The strength of this national cohort study lies within the diverse forms of mental health self-care evaluated, which were investigated by pre-existing health factors, in addition to employment status, age and gender. Findings uncover personal characteristics associated with potential barriers to engagements with mental health supports, and self-care, during an economic crisis. One limitation is that the study findings fall exclusively within the COVID-19 pandemic context, without reference to the degree of mental health self-care prior to the pandemic. However, given the declined mental health observed in this group compared to pre-pandemic reference levels (Griffiths et al. 2022b; Griffiths et al. 2021), mental health self-care would be anticipated to be less common before the pandemic when taking into context the study findings. Whilst our dataset consists of working-age Australians that are an overrepresented sample of people that experienced pandemic-related work loss, findings are valid within the context of the early stages of the pandemic in Australia, during a heightened period of work loss (ABS 2020), although they may differ based on changes in public health orders over time, and their relevance for the workforce. Within the category of formal forms of mental healthcare, the heightened use of mental health services in Australia has been ongoing since the start of the COVID-19 pandemic (AIHW 2021), demonstrating the long-term need for such services. Analogous information on mental health self-care over longer time period since the start of the pandemic remains a challenge to quantify. Longitudinal research studies, over several years, will uncover changes in informal forms of mental health self-care following periods of crisis. In many nations, stringent pandemic measures have since lifted, and unenforced recommendations remain in community settings which may promote good mental health as observed following the release of lockdown measures (Griffiths et al. 2022a). However, some workplaces remain high infection risk settings such as in the health and care industry, and mental health consequences of pandemic-related changes in work may differ from industries with fewer ongoing pandemic-related measures or changes in workload. Employers are uniquely placed to develop relevant mental health supports and policies during changes to the workplace and the nature of work resulting from the pandemic.

## Conclusion

Behavioural actions to manage mental health concerns during the COVID-19 pandemic were common, as were conversations with friends or family members. Interventions to encourage the use of formal mental health supports are encouraged such as promoting accessible mental health supports in the workplace, and increasing mental health education for all workers. Supports and services should focus on reducing barriers to health care, particularly for people associated with commonly taking less help-seeking behaviour in general, and importantly for people experiencing high levels of distress. This should be coupled with programs for mental health promotion and preventive strategies..

## Data Availability

Procedures to access data from this study are available through contacting the lead author. Proposals for collaborative analyses will be considered by the study's investigator team.

## Acknowledgments

We acknowledge the Social Research Centre for undertaking telephone interviews. We thank the participants for taking the time to complete the surveys.

## Funding

Funding was provided by Monash University and the icare Foundation. The views expressed are those of the authors and may not reflect the views of study funders. Professor Alex Collie is supported by an ARC Future Fellowship (FT190100218). The funders had no role in study design, data collection and analysis, decision to publish, or preparation of the manuscript.

## Competing interests

The authors have declared that no competing interests exist.

## Notes

### Competing Interest Statement

The authors have declared no competing interest.

### Funding Statement

Funding was provided by Monash University and the icare Foundation. Professor Alex Collie is supported by an Australian Research Council Future Fellowship (#FT190100218).

### Author Declarations

Ethical approval to conduct the study was provided by Monash University Human Research Ethics Committee (#24003).

