## Appendices for "Mental health self-care during the COVID-19 pandemic: A prospective cohort study in Australia"

**Appendix A. Actions to self-manage mental health concerns during the COVID-19 pandemic.**

| **Adjusted Odds Ratio**  **[95% Confidence Interval]** | **Lifestyle changes** | **Diet and exercise** | **Alcohol or drug use** | **Medications** | **Helplines and online resources** |
| --- | --- | --- | --- | --- | --- |
| ***Workers reporting action N (%)*** | ***1263 (78%)*** | ***764 (47%)*** | ***441 (27%)*** | ***357 (22%)*** | ***328 (20%)*** |
| ***Gender*** |  |  |  |  |  |
| Female | **2.33*** [1.82, 3.03] | **1.54*** [1.23, 1.92] | 0.92 [0.71, 1.16] | 1.27 [0.94, 1.69] | **1.56*** [1.18, 2.08] |
| Male | 1.00 (ref.) | 1.00 (ref.) | 1.00 (ref.) | 1.00 (ref.) | 1.00 (ref.) |
| ***Age Group*** |  |  |  |  |  |
| 18-24 year | 1.54 [0.87, 2.72] | **1.54*** [1.00, 2.37] | **0.48*** [0.29, 0.78] | 1.02 [0.59, 1.77] | 1.07 [0.65, 1.76] |
| 25-34 year | 1.14 [0.74, 1.74] | 0.99 [0.71, 1.40] | 0.70 [0.48, 1.01] | 0.86 [0.54, 1.36] | 1.25 [0.83, 1.87] |
| 45-54 years | 1.25 [0.86, 1.81] | 0.94 [0.69, 1.27] | 0.79 [0.57, 1.10] | 1.08 [0.72, 1.63] | 0.81 [0.55, 1.18] |
| 55-64 years | 1.04 [0.73, 1.49] | **0.73*** [0.54, 0.99] | **0.49*** [0.35, 0.69] | **1.59*** [1.07, 2.37] | 0.69 [0.47, 1.03] |
| 65+ year | 1.01 [0.58, 1.75] | **0.43*** [0.26, 0.71] | **0.41*** [0.22, 0.75] | 1.04 [0.52, 2.08] | **0.28*** [0.12, 0.69] |
| 35-44 years | 1.00 (ref.) | 1.00 (ref.) | 1.00 (ref.) | 1.00 (ref.) | 1.00 (ref.) |
| ***Employment*** |  |  |  |  |  |
| Unemployed | 0.71 [0.45, 1.11] | 0.83 [0.58, 1.19] | 0.93 [0.63, 1.37] | 0.93 [0.61, 1.42] | 0.81 [0.54, 1.22] |
| Employed | 1.00 (ref.) | 1.00 (ref.) | 1.00 (ref.) | 1.00 (ref.) | 1.00 (ref.) |
| ***Current work status*** |  |  |  |  |  |
| Not Working | **1.54*** [1.04, 2.28] | 1.35 [0.99, 1.84] | 1.09 [0.78, 1.53] | 1.14 [0.78, 1.66] | **1.79*** [1.24, 2.59] |
| Working | 1.00 (ref.) | 1.00 (ref.) | 1.00 (ref.) | 1.00 (ref.) | 1.00 (ref.) |
| ***Pre-existing anxiety*** |  |  |  |  |  |
| Anxiety | **1.59*** [1.02, 2.49] | 1.01 [0.72, 1.43] | 1.09 [0.76, 1.56] | **1.73*** [1.19, 2.51] | **1.66*** [1.13, 2.44] |
| No anxiety | 1.00 (ref.) | 1.00 (ref.) | 1.00 (ref.) | 1.00 (ref.) | 1.00 (ref.) |
| ***Pre-existing depression*** |  |  |  |  |  |
| Depression | 1.11 [0.75, 1.65] | **1.46*** [1.06, 1.99] | **1.37*** [0.98, 1.92] | **3.93*** [2.81, 5.50] | **1.58*** [1.10, 2.26] |
| No Depression | 1.00 (ref.) | 1.00 (ref.) | 1.00 (ref.) | 1.00 (ref.) | 1.00 (ref.) |
| ***Psychological distress*** |  |  |  |  |  |
| No (Low to None) | 1.00 (ref.) | 1.00 (ref.) | 1.00 (ref.) | 1.00 (ref.) | 1.00 (ref.) |
| Yes (High to Moderate) | **1.89*** [1.43, 2.56] | **1.23*** [0.99, 1.56] | **1.54*** [1.19, 2.00] | **2.63*** [1.92, 3.57] | **2.13*** [1.56, 2.86] |

*p<0.05 (bold). Models adjusted for survey mode (online or telephone).

**Appendix B. Talking about mental health concerns during the COVID-19 pandemic.**

| **Adjusted Odds Ratio**  **[95% Confidence Interval]** | **Spoken to health professional** | **Spoken to non-health professional** | **Spoken to no one** |
| --- | --- | --- | --- |
| ***Workers reporting action N (%)*** | ***303 (19%)*** | ***848 (52%)*** | ***719 (44%)*** |
| ***Gender*** |  |  |  |
| Female | 1.35 [1.00, 1.82] | **1.85*** [1.47, 2.33] | **0.55*** [0.43, 0.69] |
| Male | 1.00 (ref.) | 1.00 (ref.) | 1.00 (ref.) |
| ***Age Group*** |  |  |  |
| 18-24 year | 1.08 [0.62, 1.90] | **2.39*** [1.46, 3.90] | **0.37*** [0.22, 1.62] |
| 25-34 year | 0.87 [0.54, 1.40] | **1.47*** [1.02, 2.13] | **0.64*** [0.44, 0.94] |
| 45-54 years | 1.40 [0.92, 2.13] | 0.87 [0.63, 1.19] | 1.07 [0.77, 1.49] |
| 55-64 years | 1.26 [0.82, 1.93] | **0.56*** [0.41, 0.77] | **1.57*** [1.13, 2.18] |
| 65+ year | 0.99 [0.47, 2.08] | **0.38*** [0.23, 0.65] | **2.51*** [1.48, 4.26] |
| 35-44 years | 1.00 (ref.) | 1.00 (ref.) | 1.00 (ref.) |
| ***Employment*** |  |  |  |
| Unemployed | 1.18 [0.76, 1.83] | 0.90 [0.61, 1.33] | 1.08 [0.72, 1.63] |
| Employed | 1.00 (ref.) | 1.00 (ref.) | 1.00 (ref.) |
| ***Current work status*** |  |  |  |
| Working | 1.00 (ref.) | 1.00 (ref.) | 1.00 (ref.) |
| Not Working | 1.39 [0.94, 2.08] | **1.40*** [1.00, 1.96] | 0.72 [0.51, 1.02] |
| ***Pre-existing anxiety*** |  |  |  |
| Anxiety | **2.32*** [1.56, 3.43] | 1.22 [0.84, 1.77] | **0.50*** [0.33, 0.76] |
| No anxiety | 1.00 (ref.) | 1.00 (ref.) | 1.00 (ref.) |
| ***Pre-existing depression*** |  |  |  |
| Depression | **2.72*** [1.90, 3.90] | **1.57*** [1.12, 2.20] | **0.49*** [0.34, 0.71] |
| No Depression | 1.00 (ref.) | 1.00 (ref.) | 1.00 (ref.) |
| ***Psychological distress*** |  |  |  |
| Low to None | 1.00 (ref.) | 1.00 (ref.) | 1.00 (ref.) |
| High to Moderate | **3.45*** [2.50, 4.76] | **2.63*** [2.04, 3.33] | **0.35*** [0.28, 0.45] |

*p<0.05 (bold). Models adjusted for survey mode (online vs. telephone).

**Appendix C. Cohort profile of demographic, health, and employment characteristics.**

|  | **Cohort N (%)** |
| --- | --- |
| ***Gender*** |  |
| Female | 976 (59.3) |
| Male | 666 (40.5) |
| ***Age Group*** |  |
| 18-24 year | 114 (11.9) |
| 25-34 year | 193 (20.2) |
| 45-54 years | 417 (25.3) |
| 55-64 years | 451 (27.4) |
| 65+ year | 100 (6.1) |
| 35-44 years | 298 (18.1) |
| ***Current Employment*** |  |
| Unemployed | 274 (16.6) |
| Employed | 1372 (83.4) |
| ***Current work status*** |  |
| Working | 664 (39.1) |
| Not Working | 1002 (60.9) |
| ***Pre-existing anxiety*** |  |
| Anxiety | 266 (16.2) |
| No anxiety | 1380 (83.8) |
| ***Pre-existing depression*** |  |
| Depression | 312 (19.0) |
| No Depression | 1334 (81.0) |
| ***Current Psychological distress*** |  |
| Low to None | 855 (51.9) |
| High to Moderate | 776 (47.1) |
